# Inconsistent Rebound of Invasive Pneumococcal Disease in Connecticut following the Initial Phase of the SARS-CoV-2 Pandemic

**DOI:** 10.1101/2024.11.02.24316649

**Authors:** Stephanie Perniciaro, Daniel M. Weinberger

**Affiliations:** Department of Epidemiology of Microbial Diseases, Yale School of Public Health

## Abstract

The incidence of invasive pneumococcal disease (IPD) decreased during the SARS-CoV-2 pandemic and rebounded inconsistently over 2 years, with occasional returns to pre-pandemic levels followed by subsequent declines. We evaluated several explanations including changes in rates of viral infections that could interact with pneumococcus and changes in blood culture practices.

## Introduction

*Streptococcus pneumoniae*, the pneumococcus, is an endemic, seasonal pathogen, causing a variety of illnesses, including severe diseases like sepsis, bacteremic pneumonia, and meningitis. Invasive pneumococcal disease (IPD) is a general term for severe pneumococcal illness, generally defined as *Streptococcus pneumoniae* being isolated from a normally sterile body site (e.g. blood, cerebrospinal fluid). IPD cases in Connecticut are reported as part of the Active Bacterial Core Surveillance system (ABCs), a project of the Emerging Infections Program network coordinated by the Centers for Disease Control and Prevention (CDC).

Early in the SARS-CoV-2 pandemic, infectious disease surveillance programs in many parts of the world reported atypical patterns in IPD, with sharp decreases in the reported rates of IPD coinciding with the onset of non-pharmaceutical interventions against COVID-19 (1). Many IPD surveillance programs reported a rebound to baseline rates of IPD sometime in 2021 (2–4). It was initially believed that these declines were related to decreases in rates of exposure to the bacteria (1). However, carriage rates were largely consistent through the first period of the pandemic (5,6).

There are several possible hypotheses for the post-COVID patterns of IPD. The risk of IPD greatly increases following exposure to certain seasonal respiratory viruses (e.g., respiratory syncytial virus, influenza, human metapneumovirus). These viruses largely disappeared from circulation for the first year of the pandemic in the Northern Hemisphere, potentially influencing rates of pneumococcal disease (6–8). It is also possible that changes in healthcare seeking or use of clinical diagnostics could have influenced reported rates of infection (9).

Here we investigate trends in rates of IPD in Connecticut during the SARS-CoV-2 pandemic, 2020-2022, and explore possible explanations for the unusual dynamics during the post-COVID period.

## Methods

Hospitals and clinical microbiology laboratories reported cases of IPD to the Connecticut Department of Public Health (DPH). Serotyping by whole genome sequencing was done at the CDC’s national Streptococcal laboratory. All IPD cases in Connecticut (reviewed as exempt by the DPH Human Investigations Committee, Protocol #150E) from 2016– 2022 were included in this analysis. IPD cases were grouped by serotype (serotypes included in the 13-valent pneumococcal conjugate vaccine (PCV13, serotypes 1, 3, 4, 5, 6A, 6B, 7F, 9V, 14, 18C, 19A, 19F, 23F) or not, and by age (age <6, <18, 18-39, 18-64, ≥65 years). Complete serotype data were publicly available through 2021 for IPD cases in the other ABC sites in the United States (10), and complete serotype data were available for IPD cases in Connecticut through the end of 2022. We compared monthly IPD cases from 2020–2022 to baseline years 2016–2019 and compared serotype distributions to national data from the ABC surveillance system. Data about respiratory syncytial virus (RSV) hospitalizations in Connecticut were obtained from the CDC’s surveillance dashboard RSV-NET (11), and data about influenza hospitalizations in Connecticut were obtained from the CDC’s surveillance dashboard FluSurv-NET (12). The surveillance system for RSV hospitalizations was active in Connecticut starting in 2018. Data about COVID-19 hospitalizations were taken from the COVID-19 Reported Patient Impact and Hospital Capacity by State time series (13). We compared changes in IPD during 2020–2022 to RSV hospitalizations, influenza hospitalizations, and COVID-19 hospitalizations. State-level adherence to non-pharmaceutical interventions (NPIs) was measured with the Oxford Covid-19 Government Response Tracker (14).

Guidelines discouraging the use of blood cultures for pneumonia patients came into effect in 2020 (15). In order to determine the impact of this guidance on local blood culturing practices, data for hospitalized pneumonia cases was obtained from the largest hospital system in Connecticut. For cases of hospitalized pneumonia (as classified by the ICD-10 codes in **Supplementary Table 1**) in adults, we received a report of whether a patient had received a blood culture during their hospitalization for pneumonia. We compared the trends over time and assessed potential correlations with the rate of IPD in Connecticut. Use of the data from the large hospital system was approved by the institutional review board at Yale School of Medicine (IRB #2000036472)

## Results

1489 cases of IPD were reported in Connecticut from 2016–2022. IPD case counts in 2020, 2021 and 2022 remained below pre-pandemic baseline values, and typical seasonality was disrupted (**Figure 1**). Following the initial decline in rates of IPD starting in the Spring of 2020, rates began to increase in mid-2021. The increases were uneven in timing, with occasional increases to the expected seasonal levels followed by subsequent declines (**Figure 1**). The return to baseline IPD levels varied by age group, with younger adults (18–39 years old) surpassing baseline levels in mid-2022 (**Supplementary Figure 1**).

**Figure 1.**
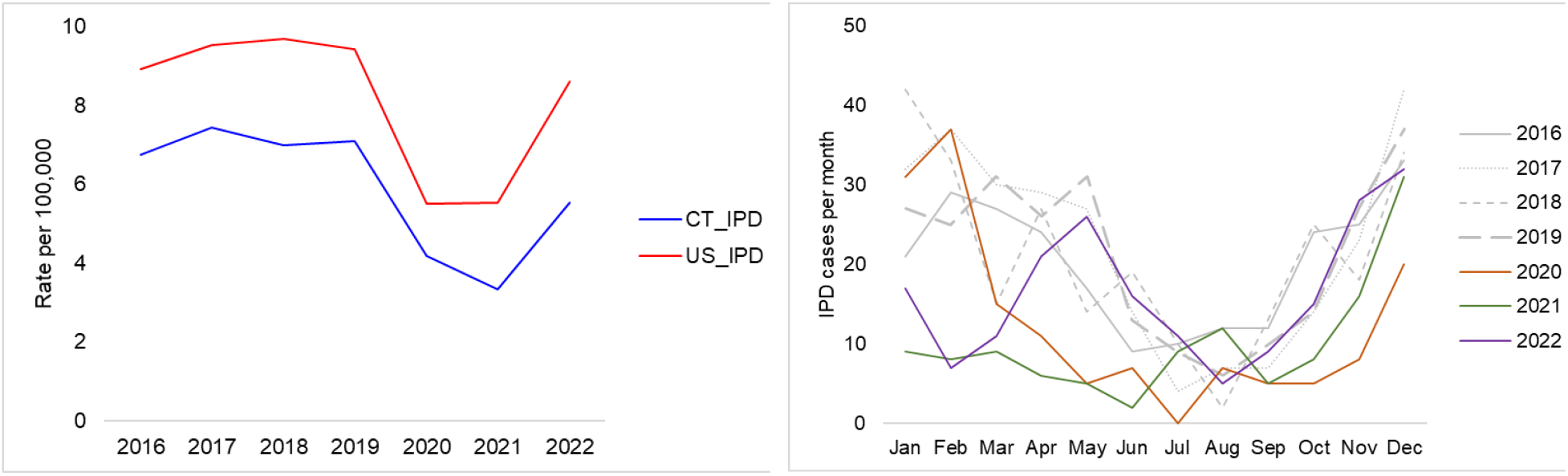
1A. Annual rates of invasive pneumococcal disease in Connecticut and in other ABC surveillance sites in the United States. 1B. Monthly rates of invasive pneumococcal disease in Connecticut, 2016-2022.

In 2016–2019, IPD cases in Connecticut typically comprised 8% of IPD in the ABC sites while in 2021 this dropped to 6.4% and in 2022 it was 6.8%. The distribution of PCV13 serotypes causing IPD in Connecticut did not change substantially following the onset of the SARS-CoV-2 pandemic (**Table 1**). In 2022, there were 22% fewer IPD cases in Connecticut than prior to the pandemic, compared to 8% fewer in the remainder of the ABC sites. There was an initial increase in the proportion of isolates that were not available for serotyping following the onset of the pandemic.

**Table 1.** Percentage of serotyped invasive pneumococcal disease isolates in Connecticut and in other surveillance sites, and percentage of 13-valent pneumococcal conjugate vaccine (PCV13) serotypes occurring from 2016-2022.

| Connecticut |  |  |  |  |  |  |  | ABC Sites |  |  |  |  |  |  |  |
| --- | --- | --- | --- | --- | --- | --- | --- | --- | --- | --- | --- | --- | --- | --- | --- |
|  | 2016 | 2017 | 2018 | 2019 | 2020 | 2021 | 2022 |  | 2016 | 2017 | 2018 | 2019 | 2020 | 2021 | 2022 |
| Total | 243 | 268 | 252 | 256 | 151 | 120 | 198 | Total | 2794 | 2988 | 3039 | 2956 | 1731 | 1736 | 2702 |
| % serotyped | 93% | 90% | 91% | 91% | 93% | 83% | 90% | % serotyped | 90% | 90% | 89% | 89% | 84% | 76% | -- |
| % PCV13 | 23% | 26% | 23% | 24% | 25% | 22% | 23% | % PCV13 | 24% | 26% | 26% | 26% | 27% | 27% | -- |

We explored several possible explanations for the decline in reported rates of IPD. First, blood culture is used to detect pneumococcus and is needed for most diagnoses of IPD. For all adults hospitalized with a primary diagnosis of pneumonia at Hospital A, blood culture rates decreased from 2020–2022, from 64% to around 52% (**Figure 2**). There were 44 fewer blood cultures performed in 2022 compared to 2020. Second, the increases in rates of IPD starting in mid-2021 roughly aligned with epidemics of influenza and RSV (**Figure 3**). Finally, average stringency in adherence to NPIs was consistent between Connecticut and other ABC surveillance sites, so that is unlikely to have contributed to differences between sites. (**Supplementary Figure 2**).

**Figure 2.**
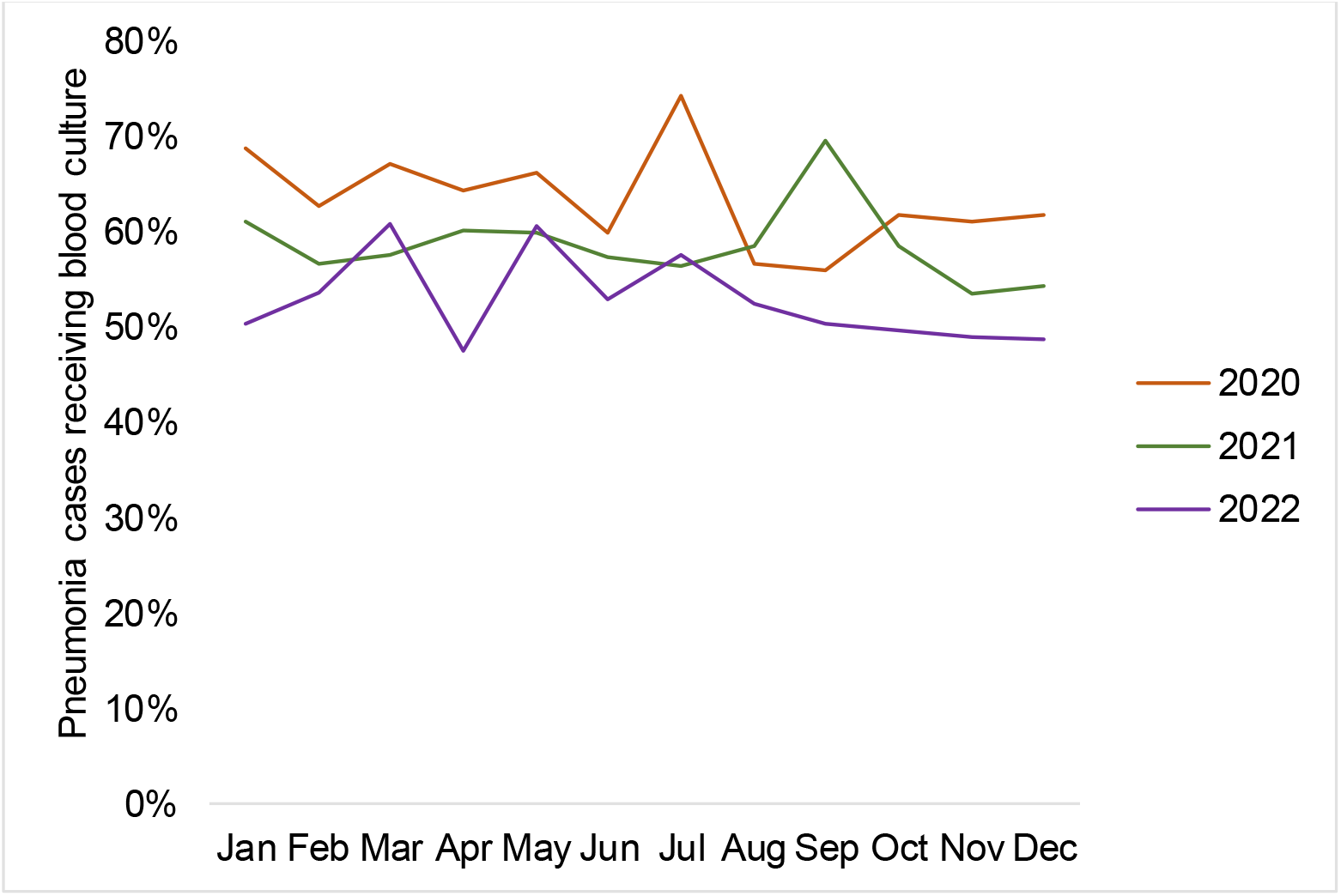
Blood culture rates for hospitalizations with a primary diagnosis of pneumonia in a large hospital system, 2020-2022.

**Figure 3.**
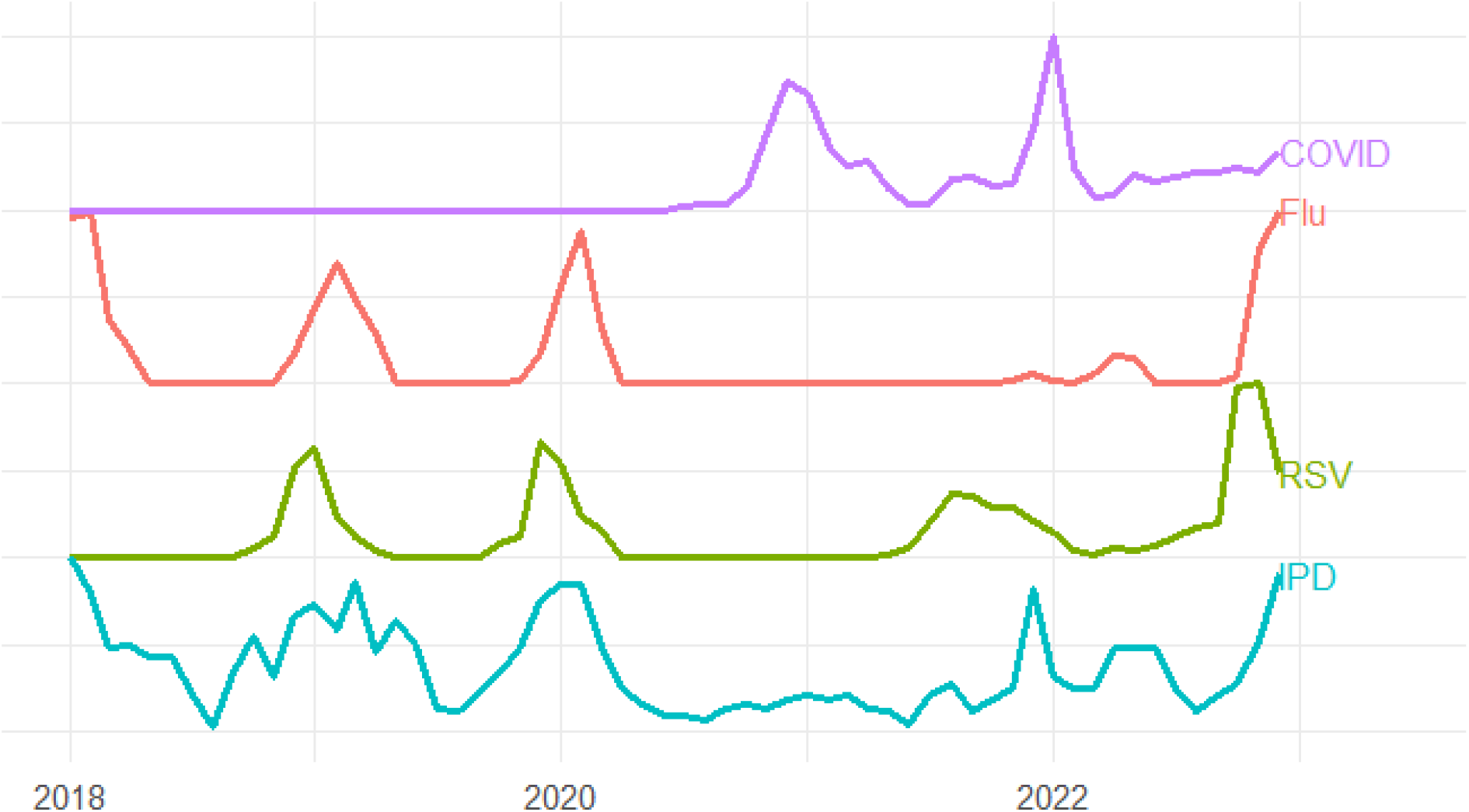
Rates of invasive pneumococcal disease, respiratory syncytial virus hospitalizations, influenza hospitalizations, and COVID-19 hospitalizations in Connecticut, 2018-2022.

## Discussion

Here we present IPD data from statewide active surveillance, which shows declines in rates of IPD that persisted through mid-2021. Rates of IPD began to recover towards typical seasonal levels in concert with epidemics of RSV and influenza. Lower rates of blood culturing could partially explain why rates of IPD remained below expected levels for parts of 2022.

No major differences in PCV13-type serotype distribution of IPD in Connecticut were noted over the study period, which suggests that despite the disruptions to pneumococcal disease epidemiology, the potential impact of pneumococcal vaccines is unchanged. The age distribution of IPD also remained relatively constant in both Connecticut and other ABCs sites. Unlike reports from other countries, IPD in Connecticut did not consistently return to baseline values, with low values persisting through parts of 2022. In August of 2021 and June–July of 2022, there were off-season increases in IPD cases. Viral surveillance data indicate that these values had returned to or surpassed pre-pandemic levels, and NPIs remained consistently low across ABC sites, so the reason for continued low case counts of IPD during intervening periods is unclear.

We examined the possibility of a change to blood culture recommendations impacting IPD discovery. The rate of blood culture for primary pneumonia diagnoses did decrease by almost 20%, and it is unclear how much this change in practice at one large hospital system contributed to reduced detection of IPD statewide.

This analysis has several limitations. Surveillance catchment areas for IPD, RSV, and influenza are slightly different, and it is possible that these differences could affect comparisons between these conditions. We investigated associations with NPIs, with viral infections, and with blood culture practices in the state’s largest hospital system, but there could be other factors influencing IPD rates in Connecticut, including reporting differences, vaccination practices, and blood culture practices in other hospital systems. Differences in reporting practices due to personnel and resource shortfalls within the healthcare system are one potential reason for this prolonged decline, but it seems unlikely that Connecticut would be more affected by these issues than other ABC sites, as ABC sites are required to verify complete case reports.

## Conclusions

Invasive pneumococcal disease in Connecticut rebounded inconsistently to pre-SARS-CoV-2-pandemic levels, hitting baselines values mid-2021 and then dipping below baseline until the second half of 2022, despite other indications that respiratory illnesses had returned to pre-pandemic levels. Hospitalizations from RSV returned to or surpassed baseline values in late 2021, and influenza hospitalizations returned to baseline values in 2022. Connecticut did not have stricter adherence to non-pharmaceutical interventions in 2021 and 2022. These findings suggest that both the elimination and resurgence of respiratory viruses as well as changes in blood culture practices could have contributed to the observed patterns.

## Data Availability

All data produced in the present study are available upon reasonable request to the authors

## Funding and Acknowledgements

This work was supported in part by a research grant from Investigator-Initiated Studies Program of Merck Sharp & Dohme Corp. The opinions expressed in this paper are those of the authors and do not necessarily represent those of Merck Sharp & Dohme Corp. This publication was made possible by CTSA grant number KL2 TR001862 from the National Center for Advancing Translational Science (NCATS), a component of the National Institutes of Health (NIH). Its contents are solely the responsibility of the authors and do not necessarily represent the official view of NIH. The authors would like to thank Susan Petit and Lynn Sosa from the Connecticut Department of Public Health for their input and assistance with the preparation of this manuscript.

## Supplementary Figures and Tables

**Supplementary Figure 1.**
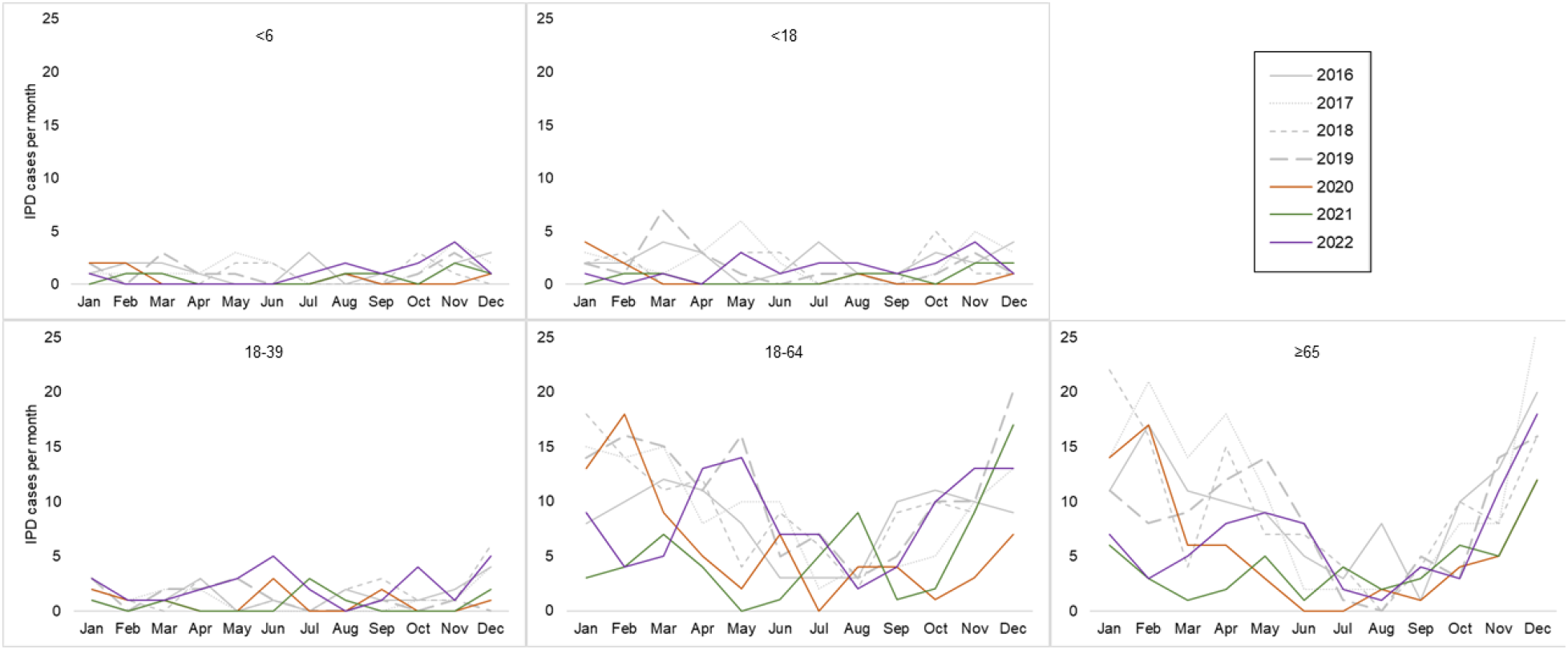
Invasive pneumococcal disease by age group in Connecticut, 2016-2022.

**Supplementary Figure 2.**
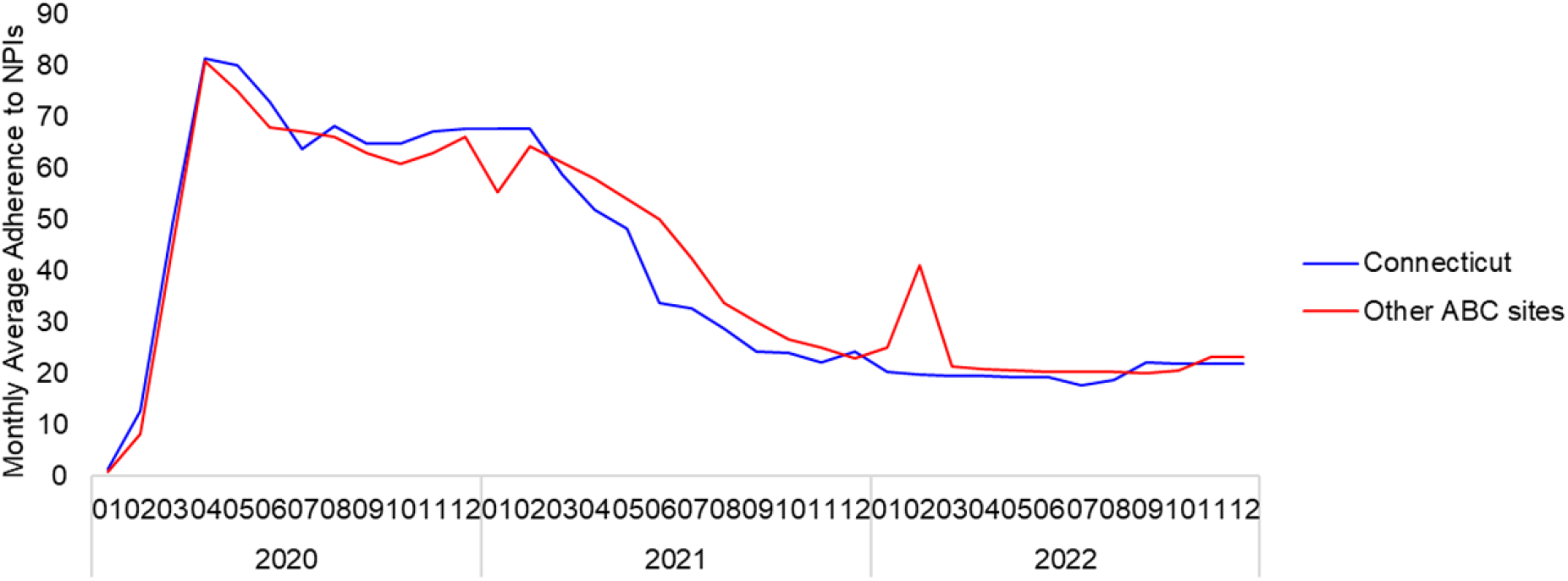
Monthly average stringency score for adherence to nonpharmaceutical interventions, Connecticut and other Active Bacterial Core surveillance sites, 2020-2022.

**Supplementary Table 1.** ICD-10 diagnostic codes associated with pneumonia, empyema, and septicemia.

|  |  |
| --- | --- |
| Pneumonia or Empyema | J10.00, J10.01, J10.08,<br>J11.00, J11.08, J12.0, J12.1,<br>J12.2, J12.81, J12.82, J12.3,<br>J12.89, J12.9, J13, J18.1,<br>J15.0, J15.1, J14, J15.4,<br>J15.3, J15.20, J15.211,<br>J15.212, J15.29, J15.5,<br>J15.6, J15.7, J15.8, J15.9,<br>J16.0, J16.8, J17, J18.0,<br>J18.8, J18.9, J86.0, J86.9,<br>A2.21, A37.01, A37.11,<br>A37.81, A37.91, A48.1,<br>B25.0, B4.40, B77.81 |
| Septicemia | A02.1, A20.7, A22.7, A26.7,<br>A31.2, A32.7, A39.2, A39.3,<br>A39.4, A40.0, A40.1, A40.3,<br>A40.8, A40.9, A41.01,<br>A41.02, A41.1, A41.2, A41.3,<br>A41.4, A41.50, A41.51,<br>A41.52, A41.53, A41.59,<br>A41.81, A41.89, A41.9,<br>A42.7, A54.86, B00.7,<br>B37.7, R65.20, R65.21,<br>R78.81 |

## References

1. Brueggemann AB, Jansen van Rensburg MJ, Shaw D, McCarthy ND, Jolley KA, Maiden MCJ, et al. Changes in the incidence of invasive disease due to Streptococcus pneumoniae, Haemophilus influenzae, and Neisseria meningitidis during the COVID-19 pandemic in 26 countries and territories in the Invasive Respiratory Infection Surveillance Initiative: a prospective analysis of surveillance data. Lancet Digit Health. 2021 Jun 1;3(6):e360–70.

2. Bertran M, Amin-Chowdhury Z, Sheppard CL, Eletu S, Zamarreño DV, Ramsay ME, et al. Increased Incidence of Invasive Pneumococcal Disease among Children after COVID-19 Pandemic, England - Volume 28, Number 8—August 2022 - Emerging Infectious Diseases journal - CDC.; Available from: https://www.nc.cdc.gov/eid/article/28/8/22-0304_article

3. Rybak A, Assad Z, Levy C, Bonarcorsi S, Béchet S, Werner A, et al. Age-Specific Resurgence in Invasive Pneumococcal Disease Incidence in the COVID-19 Pandemic Era and Its Association With Respiratory Virus and Pneumococcal Carriage Dynamics: A Time-Series Analysis. Clin Infect Dis. 2024 Apr 15;78(4):855– 9.

4. Ricketson LJ, Kellner JD. Changes in the Incidence of Invasive Pneumococcal Disease in Calgary, Canada, during the SARS-CoV-2 Pandemic 2020–2022. Microorganisms. 2023 May;11(5):1333.

5. Danino D, Ben-Shimol S, Van Der Beek BA, Givon-Lavi N, Avni YS, Greenberg D, et al. Decline in Pneumococcal Disease in Young Children during the COVID-19 Pandemic in Israel Associated with Suppression of seasonal Respiratory Viruses, despite Persistent Pneumococcal Carriage: A Prospective Cohort Study. Clin Infect Dis Off Publ Infect Dis Soc Am. 2021 Dec 14;ciab1014.

6. Willen L, Ekinci E, Cuypers L, Theeten H, Desmet S. Infant Pneumococcal Carriage in Belgium Not Affected by COVID-19 Containment Measures. Front Cell Infect Microbiol. 2021;11:825427.

7. Dagan R, Danino D, Weinberger DM. The Pneumococcus–Respiratory Virus Connection—Unexpected Lessons From the COVID-19 Pandemic. JAMA Netw Open. 2022 Jun 28;5(6):e2218966.

8. Ouldali N, Deceuninck G, Lefebvre B, Gilca R, Quach C, Brousseau N, et al. Increase of invasive pneumococcal disease in children temporally associated with RSV outbreak in Quebec: a time-series analysis. Lancet Reg Health - Am. 2023 Feb 15;19:100448.

9. Dirkx KKT, Mulder B, Post AS, Rutten MH, Swanink CMA, Wertheim HFL, et al. The drop in reported invasive pneumococcal disease among adults during the first COVID-19 wave in the Netherlands explained. Int J Infect Dis. 2021 Oct;111:196– 203.

10. 1998-2021 Serotype Data for Invasive Pneumococcal Disease Cases by Age Group from Active Bacterial Core surveillance | Data | Centers for Disease Control and Prevention. Available from: https://data.cdc.gov/Public-Health-Surveillance/1998-2021-Serotype-Data-for-Invasive-Pneumococcal-/qvzb-qs6p/about_data

11. RSV-NET Interactive Dashboard | CDC. Available from: https://www.cdc.gov/rsv/research/rsv-net/dashboard.html

12. Influenza Hospitalization Surveillance Network (FluSurv-NET) | CDC. Available from: https://www.cdc.gov/flu/weekly/influenza-hospitalization-surveillance.htm

13. COVID-19 Reported Patient Impact and Hospital Capacity by State Timeseries (RAW) | HealthData.gov. Available from: https://healthdata.gov/Hospital/COVID-19-Reported-Patient-Impact-and-Hospital-Capa/g62h-syeh/about_data

14. Hale T, Webster S, Petherick A, Phillips T, Kira B. Oxford COVID-19 Government Response Tracker, Blavatnik School of Government. Available from: https://covidtracker.bsg.ox.ac.uk/

15. Metlay JP, Waterer GW, Long AC, Anzueto A, Brozek J, Crothers K, Cooley LA, Dean NC, Fine MJ, Flanders SA, Griffin MR, Metersky ML, Musher DM, Restrepo MI, Whitney CG; on behalf of the American Thoracic Society and Infectious Diseases Society of. Diagnosis and Treatment of Adults with Community-acquired Pneumonia. An Official Clinical Practice Guideline of the American Thoracic Society and Infectious Diseases Society of America. Available from: https://www.idsociety.org/practice-guideline/community-acquired-pneumonia-cap-in-adults/

